# Emergency Department Quality of Care for Sickle Cell Disease in Ontario, Canada: A Population-Based Matched Cohort Study

**DOI:** 10.1101/2022.07.06.22277294

**Authors:** Derek C. H. Chan, Fiona G. Kouyoumdjian, Andrea J. Pang, Yue Niu, Uma H. Athale, Jacob Pendergrast, Madeleine M. Verhovsek

**Affiliations:** Michael G. DeGroote School of Medicine, Faculty of Health Sciences, McMaster University, Hamilton, Ontario, Canada; Department of Pediatrics, British Columbia Children’s Hospital, Faculty of Medicine, University of British Columbia, Vancouver, British Columbia, Canada; Department of Family Medicine, Faculty of Health Sciences, McMaster University, Hamilton, Ontario, Canada; Populations and Public Health Research Program, ICES Central, Toronto, Ontario, Canada; McMaster Children’s Hospital, Hamilton, Ontario, Canada; Division of Pediatric Hematology/Oncology, Department of Pediatrics, Faculty of Health Sciences, McMaster University, Hamilton, Ontario, Canada; Laboratory Medicine Program, Toronto General Hospital, University Health Network, Toronto, Ontario, Canada; Department of Laboratory Medicine and Pathobiology, University of Toronto, Toronto, Ontario, Canada; St. Joseph’s Hospital, Hamilton, Ontario, Canada; Division of Hematology and Thromboembolism, Department of Medicine, Faculty of Health Sciences, McMaster University, Hamilton, Ontario, Canada

**Author notes:** **Corresponding Author:** Derek Chan.

## Abstract

**Importance:** Acute vaso-occlusive crisis (VOC) in sickle cell disease (SCD) often leads patients to seek emergency department (ED) care. The landscape of SCD ED care within Canada’s universal and publicly funded healthcare system remains largely undefined.

**Objective:** To characterize the quality of SCD VOC ED and inpatient care in Ontario, Canada.

**Design, Setting and Participants:** We used population-level health administrative data to identify patients with SCD in Ontario, Canada who presented to the ED with SCD VOC (ICD-10-CA code D57.0) from 2006 to 2018. We derived two reference patient cohorts from the general population and separately from a validated Crohn’s disease cohort matched at the level of ED visits by age, sex, neighbourhood income quintile, geography, date range of visit, and ED triage score.

**Main Outcomes and Measures:** Quality of care metrics included the number of ED visits, triage scores, time to physician initial assessment, time from triage to disposition, type of disposition, number of hospital admissions, length of admission for those admitted to hospital, number of repeat ED visits within a 14- and 30-day time window, and number of ED visits within any 1-year period. Stratifying variables and sociodemographic data included the month and year of ED visit, age bracket, triage scores, sex, rurality, Ontario’s marginalization indices (neighbourhood income, dependency, deprivation, ethnic concentration, instability), and number of ED visits within any 1-year period.

**Results:** We identified 2,123 patients who presented to the ED with SCD VOC for a total of 18,712 unscheduled ED visits. Annual ED visit rates ranged from a mean of 2.6 to 14.2 visits per patient, with 4.6% of patients having incurred 38.4% of all SCD VOC ED visits. Subgroup analyses showed that adult patients with SCD VOC visited the ED 2.5 times more than pediatric patients, but their wait times for initial physician assessment were significantly longer (90.6 vs. 51.1 minutes, *p* < 0.0001) with a difference that was disproportionately larger than the reference cohorts. Males with SCD VOC experienced significantly shorter ED and inpatient stays compared to females (e.g. admission length of stay: 4.4 vs. 5.8 days, *p* < 0.0001), but their ED return rates within 30-days were significantly higher (0.89 vs. 0.35 times, *p* < 0.0001) with a difference that was disproportionately larger relative to the reference cohorts. Lower-income neighbourhoods correlated with lower acuity triage scores for SCD VOC visits and increased repeat ED visit rates at more pronounced levels than the reference cohorts. Higher acuity triage scores for adults with SCD VOC correlated with improved metrics for quality of ED and inpatient care with fewer longer-term ED visits.

**Conclusions and Relevance:** Patients with SCD VOC carry a significant burden of acute care needs, with a smaller patient subset whose high ED visit rates have distinctly increased over time in Ontario, Canada. Adults, males and those from lower income quintile neighbourhoods with SCD VOC experienced lower quality of care metrics compared to their counterparts, with differences that were disproportionately larger than those found in the general ED population and in patients with Crohn’s disease. Higher acuity ED triage scores reflected improved downstream quality of care metrics for adult patients with SCD VOC. Future work should address these identified care quality disparities for SCD VOC that remain present within a universal and publicly funded healthcare system.

**Key Points:** *Question:* What is the quality of emergency department (ED) and inpatient care for patients with sickle cell disease (SCD) vaso-occlusive crisis (VOC) in Ontario, Canada?

*Findings:* This population-based matched cohort study of 2,123 patients with 18,712 ED visits for SCD VOC identified that adults, males and those from lower-income neighbourhoods experienced lower quality of care metrics compared to their counterparts, with differences that were disproportionately larger than those in two reference patient cohorts.

*Meaning:* Disparities in the quality of SCD VOC care with their associated outcomes remain pronounced and present within a universal and publicly funded healthcare system.

## Introduction

Sickle cell disease (SCD) is an inherited haemoglobin disorder that is associated with chronic and life-threatening complications and periodically presents with painful vaso-occlusive crises (VOCs). This disease predominantly occurs in people of African, Mediterranean, Middle Eastern and/or Indian ancestry. The acute, unpredictable and debilitating nature of SCD VOCs frequently leads patients to seek emergency department (ED) care, where the quality of care provided within two-tiered healthcare settings is known to be often inconsistent and poor.^1–7^ For the estimated 5,000 Canadians who have SCD, more than 50% of the affected population reside in the province of Ontario.^8,9^ Little is known about their quality of ED and inpatient care within Canada, where health care is universal and publicly funded.

Current data on clinical care for Canadians with SCD VOC remain limited. Among the few published studies, all have been limited to single hospital settings. Previous work has shown that the incidence of paediatric SCD VOC correlates with colder temperatures and higher wind speed,^10^ with a higher chance of hospital admission predicted by a higher acuity pain score at ED triage, older age and increased systolic blood pressure.^11^ Early work among adults with SCD VOC has identified potential variability in ED triage scoring^12^ and time incurred before patients receive opioid administration for acute pain,^13^ leading to a recent trial of an alternative short-stay unit model of care^14^ as a means to explore ways to close such gaps of inconsistent care.

To characterize the quality of SCD VOC ED and inpatient care provided across Ontario, we pursued a population-level analysis of health administrative datasets in which SCD VOC ED visit rates, triage scores, time to physician assessment, ED disposition, duration of hospital stay and repeat ED visit rates were measured, stratified by various sociodemographic variables, and compared to the general ED patient population and to patients with Crohn’s disease, with the latter group selected on the basis of having an acute-on-chronic course with variable pain episodes leading both pediatric and adult patients from different ethnic backgrounds to seek acute ED care.

## Methods

### Study settings

Between 2006 and 2018, the number of ED facilities in Ontario, Canada increased from 171 to 178. Mandated reporting of ED data into the National Ambulatory Care Reporting System (NACRS) and hospital data into the Discharge Abstract Database (DAD) resulted in a full reporting compliance rate (apart from 4 facilities excluded in 2006 due to data quality issues). EDs and hospital healthcare services in Ontario are universal and publicly funded for all Canadian citizens and permanent residents through the Ontario Health Insurance Plan (OHIP) while refugee claimants are insured through the Interim Federal Health Program. Canadians from outside of Ontario have health care fees charged to their home province’s public insurance program. Private health care insurance is available to non-Canadian citizens. Individuals lacking any insurance coverage are charged directly.

### Patient cohort definitions and matching criteria

We queried NACRS and DAD with the ICD-10-CA code D57.0 (Sickle-cell anaemia with crisis) under admitting diagnosis and most responsible diagnosis to identify all patients who presented to the ED with a primary SCD VOC between 2006 to 2018, limited to unscheduled ED visits and to patients with OHIP. These datasets were linked using unique encoded identifiers and analyzed at ICES, which is an independent, non-profit research institute whose legal status under Ontario’s health information privacy law allows it to collect and analyze health care and demographic data, without consent, for health system evaluation and improvement. We derived matched patient cohorts from the general ED patient population and separately for those with Crohn’s disease (*Figure 1A*), with ED visits from the SCD VOC cohort being matched at a 1-to-2.7 ratio to ED visits from the general ED population by age, sex, neighbourhood income quintile, geography (forward sortation area) and date range of visit (−15 to +15 days of each ED visit), and a 1-to-1.75 ratio obtained when additionally matching by ED triage score. SCD VOC ED visits were similarly matched to ED visits from a validated Crohn’s disease patient cohort at a 1-to-1.16 ratio by similar criteria except for using broader geography (census division) and date range of visit (−45 to +45 days of each ED visit) parameters, with a 1-to-1.1 ratio obtained when additionally matching by ED triage score.

**Figure 1:**
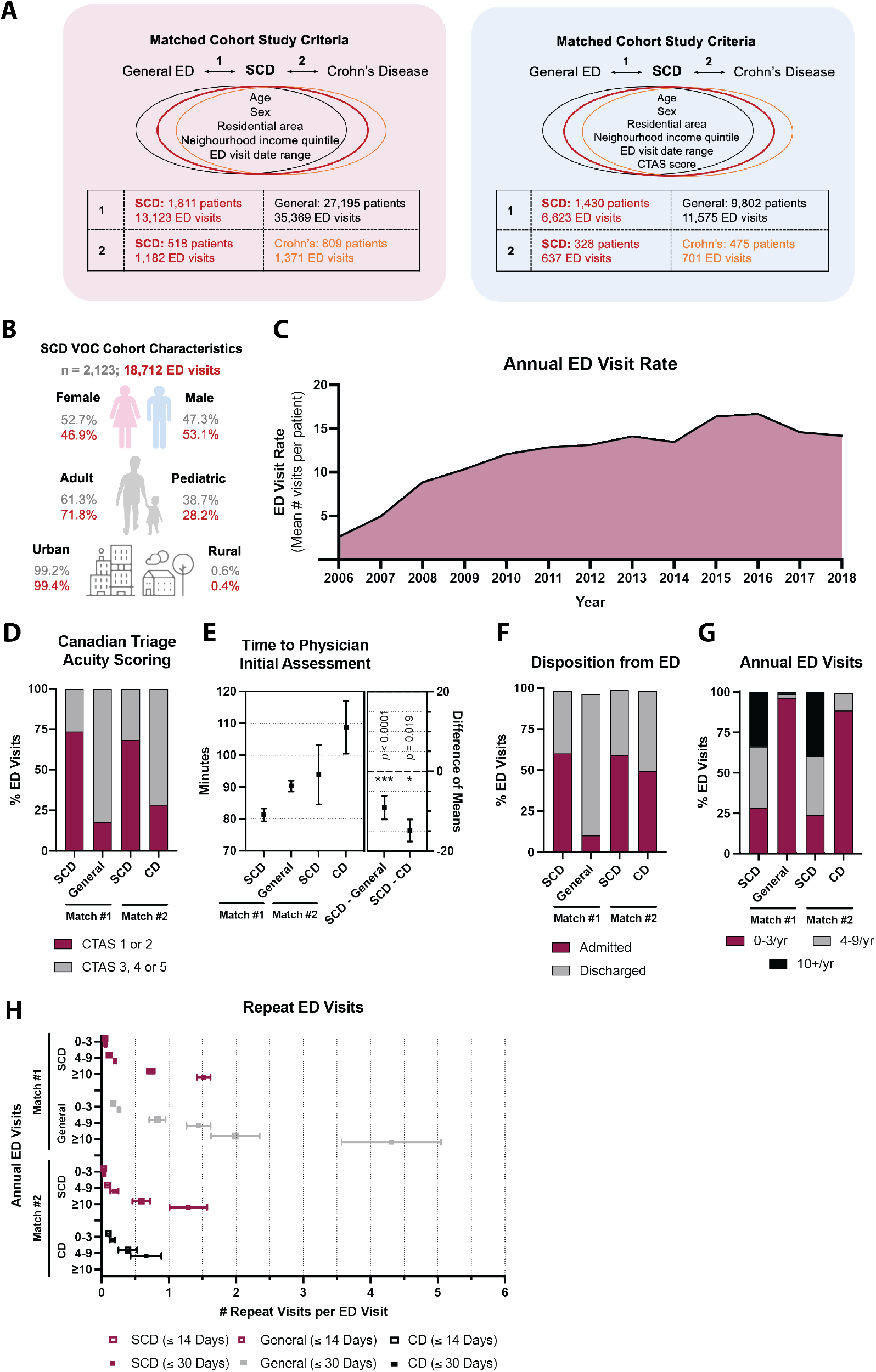
Patients with SCD VOC carry a significant burden of acute care needs. A) Matched cohort study criteria and patient numbers between SCD VOC and the general ED population (*Match #1*) and between SCD VOC and a validated Crohn’s disease cohort (*Match #2*) without (*left*) and with (*right*) additional CTAS ED triage as a criterion; B) SCD VOC cohort characteristics by sex, age range and type of residence; C) Annual SCD VOC ED visit rates; Matched cohort comparisons based on ED triage score (D), time to initial physician assessment (E), ED disposition (F), and any 1-year ED visit frequency between 0–3/yr, 4–9/yr or ≥10/yr (G); H) ED visit frequency categories stratified by repeat ED visits within 14- and 30-days; Error bars represent 95% confidence intervals (CI).

### Quality of care metrics

The following ED and hospital metric data were extracted from NACRS and DAD: number of ED visits, triage scores as per the Canadian Triage and Acuity Scale (CTAS), time to physician initial assessment, time from triage to disposition, type of disposition, number of hospital admissions, length of admission for those admitted to hospital, number of repeat ED visits within a 14- and 30-day time window, and number of ED visits within any 1-year period. Stratifying variables for extracted metric data included the following: month and year of ED visit, age bracket, CTAS scores, sex (in relation to emerging data suggestive of sex differences in SCD clinical outcomes^15^), rurality, Ontario’s marginalization indices,^16^ and number of ED visits within any 1-year period.

### Statistical analyses

Categorical sums and means with 95% confidence intervals were determined for applicable ED and hospital metric data. Differences between means were calculated at the same confidence interval threshold with *p*-values < 0.05 being considered statistically significant. Bivariate regression analyses for the SCD VOC ED patient cohort were conducted using the generalized linear method on Statistical Analysis System (SAS) software with sociodemographic and ED metric stratifiers as independent variables and the following clinically relevant outcomes as dependent variables: ED disposition (admission versus discharge), length of admitted inpatient stay, repeat ED visit within 30 days, and classification as a high ED user within a 1-year period (≥4 visits/year versus 0-3 visits/year).

## Results

### Patients with SCD VOC carry a significant burden of acute care needs, with a smaller patient subset whose high ED visit rates are distinctly increasing over time

We identified 2,123 patients who presented to the ED with SCD VOC for a total of 18,712 ED visits between January 1, 2006 and December 31, 2018, with background characteristics shown in *eTable 1*. There were more visits for adults (71.8%) than children (28.2%) and for males (53.1%) than females (46.9%) (*Figure 1B*). ED visit rates varied between a mean of 2.6 to 14.2 visits per patient per year from 2006 to 2018 (*Figure 1C*). The distribution of number of ED visits per year varied substantially across the cohort: 4.6% (*n* = 99) had ≥10 visits/year, accounting for 38.4% of all visits, while 20.6% (*n* = 437) had 4–9 visits/year, representing 36.2% of visits, and 67.0% (*n* = 1,423) had 1–3 visits/year, making up the remaining 25.4% of visits. Bivariate regression analyses between various sociodemographic variables with four selected clinically relevant outcomes (ED disposition, length of hospital admission, repeat ED visits within 30 days, any 1-year ED visit frequency) revealed varying significant degrees of association (*eTable 2*). Cohort comparisons further reflecting the significant burden of acute care needs for SCD VOC cases are found in the *Supplement*.

### Adult patients with SCD VOC have a greater burden of need and are not receiving equal or recommended ED treatment compared to pediatric patients, with differences in quality of care metrics that are disproportionately larger than the general ED population and to patients with Crohn’s disease

Subgroup analyses between adult and pediatric patients showed that annual ED visit rates for adults with SCD VOC were 2.5 to 1.5 times higher compared to pediatric patients, yet only 1.1 to 1.0 times higher in the reference cohorts (*Figure 2A*). 17.7% of adult SCD VOC ED visits were assigned disproportionately lower triage scores (CTAS 3, 4 or 5) compared to pediatric counterparts, a difference that was not observed in the general ED population, but was similar in patients with Crohn’s disease (*Figure 2B*). While the other 68.4% of adult SCD VOC ED visits were triaged at CTAS 1 or 2 compared to 18.2% in the general ED population, adult SCD VOC cases were only assessed at −4.4 minutes (95% CI −6.53–-2.33) sooner (*p* < 0.0001). After adjusting for ED triage score, adult SCD VOC cases waited on average 90.6 (95% CI 88.1–93.1) to 96.8 minutes (95% CI 87.0–106.6) for initial physician assessment, which was 39.5 (95% CI 34.77–44.21, *p* < 0.0001) to 41.1 minutes (95% CI 4.4–77.8, *p* = 0.028) longer than pediatric patients (*Figure 2C*), placing adult patients with SCD VOC well behind clinically recommended targets of receiving first-dose analgesia within 30 minutes of triage or 60 minutes from registration.^8,12^ Similar findings were mirrored when assessing ED time between triage and disposition between adult and pediatric patients with SCD VOC compared to both reference cohorts (*Figure 2D*). 18.5 to 31.6% of adult SCD VOC cases were disproportionately discharged compared to pediatric counterparts, while only a 2 to 0.4% difference was found in the reference cohorts (*Figure 2E*). Among hospital admissions, adult SCD VOC length of stay averaged 5.8 (95% CI 5.6–6.0) to 6.1 days (95% CI 5.5–6.7), which was 2.2 (95% CI 1.8–2.7, *p* < 0.0001) to 0.66 days (95% CI −1.6–2.9, *p* = 0.58) longer than pediatric patients, while adults from the reference cohorts stayed 0.34 days (95% CI 0.01–0.67, *p* = 0.045) longer to −2.47 days shorter (95% CI −4.46–-0.48, *p* = 0.015) than pediatric patients (*Figure 2F*). Repeat ED visit rates within 30-days for SCD VOC showed more pronounced differences between adult and pediatric patients compared to both reference cohorts (*Figure 2G*). Over any 1-year period, adult patients with SCD VOC also incurred a disproportionately larger burden of ED visits, with a ≥10 visits/year frequency that did not register in either reference cohorts (*Figure 2H*).

**Figure 2:**
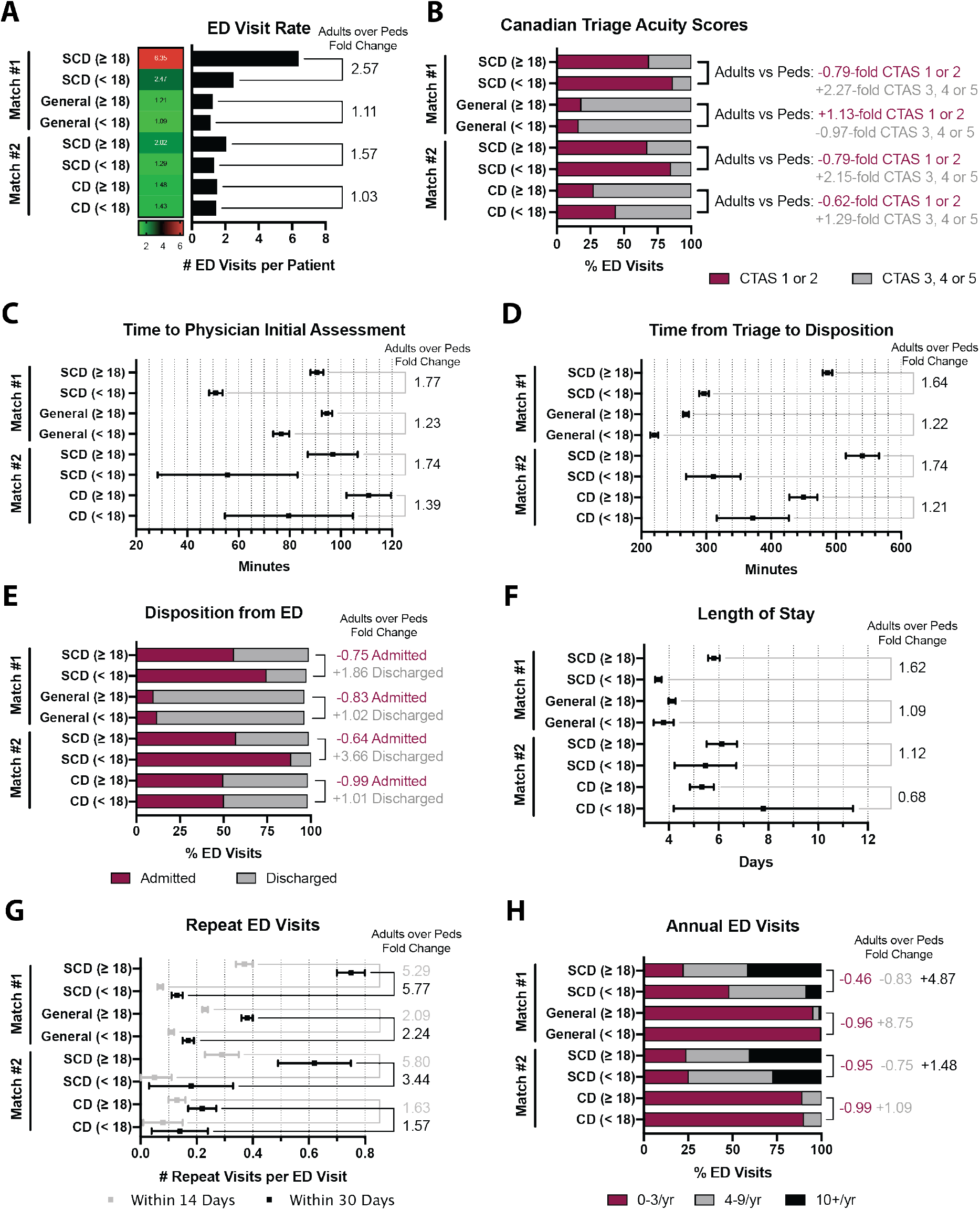
Adult patients with SCD VOC have a greater burden of need and are not receiving equal or recommended ED treatment compared to pediatric patients. Adult versus pediatric subgroup analyses for A) overall ED visit rate; B) acuity triage CTAS score; C) time to initial physician assessment; D) time in ED between triage and disposition; E) ED disposition; F) hospital admission length of stay; G) repeat ED visits within 14- and 30-days; and H) any 1-year ED visit frequency between 0–3/yr, 4–9/yr or ≥10/yr; Error bars represent 95% CI.

### Male patients presenting with SCD VOC experience shorter stays in the ED and hospital compared to female patients, but their return rates are disproportionately higher, even when compared to the general ED population and to patients with Crohn’s disease

Sex-based subgroup analyses revealed no significant differences in annual ED visit rates or triage scoring between males and females with SCD VOC. While males in all groups waited slightly less compared to females for initial physician assessment (*Figure 3A*), males with SCD VOC spent −64.5 minutes (95% CI −76.8–-52.1, *p* < 0.0001) less time between ED triage and disposition compared to females, whereas this difference was only −20.4 minutes (95% CI −27.9– −12.9) in the general ED population (*Figure 3B*). 5% of male SCD VOC ED visits were disproportionately discharged compared to females, a difference that was not found in either reference cohorts (*Figure 3C*). Among hospital admissions, males with SCD VOC experienced significantly shorter lengths of stay at 4.4 (95% CI 4.2–4.6) to 4.7 days (95% CI 4.2–5.2), which were −1.40 (95% CI −1.74–-1.06, *p* < 0.0001) to −2.45 days (95% CI −3.6–-1.3, *p* < 0.0001) shorter compared to females, differences that were either more pronounced compared to or absent in the reference cohorts (*Figure 3D*). Males with SCD VOC had a significantly higher repeat ED visit rate within 30-days compared to females with a mean difference of 0.54 visits (95% CI 0.46–0.62, *p* < 0.0001), a finding that was not observed in either reference cohorts (*Figure 3E*). Males with SCD VOC also experienced a disproportionately higher ≥10 visits/year frequency over any 1-year period with a 3.7 to 14.7% increase over females, while this visit frequency barely registered in the reference cohorts (*Figure 3F*).

**Figure 3:**
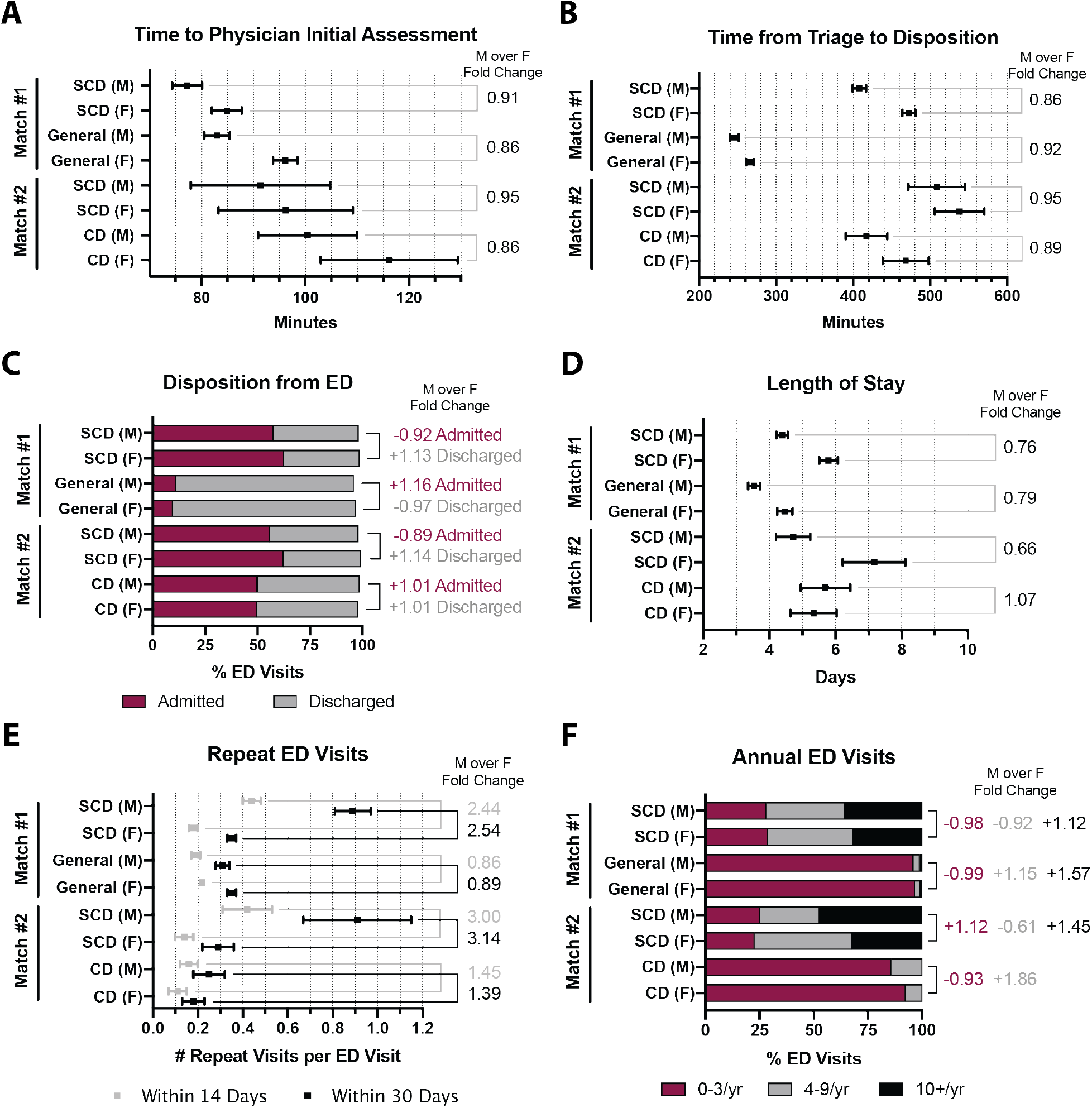
Male patients presenting with SCD VOC experience shorter stays in the ED and hospital compared to female patients, but their return rates are disproportionately higher. Male versus female subgroup analyses for A) time to initial physician assessment; B) time in ED between triage and disposition; C) ED disposition; D) hospital admission length of stay; E) repeat ED visits within 14- and 30-days; and F) any 1-year ED visit frequency between 0–3/yr, 4–9/yr or ≥10/yr; Error bars represent 95% CI.

### Lower income neighbourhood patients with SCD VOC are associated with lower acuity triage scores and increased repeat ED visits at levels that are more pronounced than the general ED population and to patients with Crohn’s disease

Subgroup analyses by neighbourhood income quintile revealed that the lowest quintile among patients with SCD VOC carried the highest ED visit rates compared to either reference cohorts (*Figure 4A*). Differences between the lowest and highest quintiles for the SCD VOC cohort were disproportionately larger compared to the general ED population, and similar in extent to patients with Crohn’s disease, although this latter comparison was limited by few patients in the highest quintiles. Lower neighbourhood income quintiles among patients with SCD VOC correlated with lower acuity triage scores (CTAS 3, 4 or 5), but this finding was not present in either reference cohorts (*Figure 4B*). No obvious patterns were identified when analyzing time to initial physician assessment, time between triage and disposition, ED disposition, and admission length of stay when these metrics were stratified by neighbourhood income quintile. Lower neighbourhood income quintiles among patients with SCD VOC reflected a higher frequency of repeat ED visits within 30-days with a mean difference of 0.64 visits (95% CI 0.40–0.88) (*p* < 0.0001) between the lowest and highest quintile, which was disproportionately larger than 0.25 visits (95% CI 0.13–0.37) (*p* < 0.0001) in the general ED population, and similar in extent to patients with Crohn’s disease, although this latter comparison was again limited by few patients in the highest quintiles. Lower neighbourhood income quintiles also correlated with a greater number of ED visits for SCD VOC over any 1-year period, a pattern that barely registered in either reference cohorts (*Figure 4D*).

**Figure 4:**
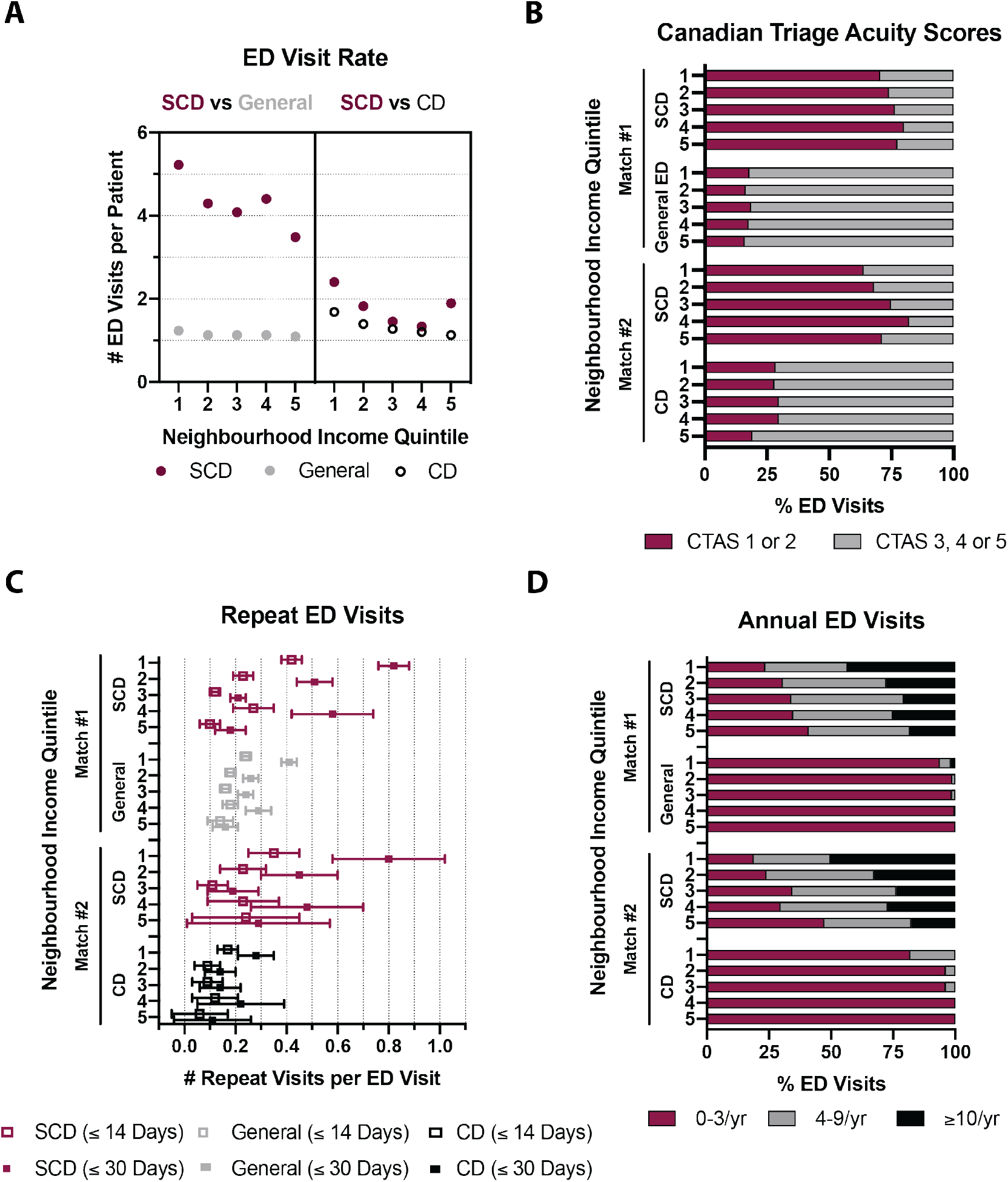
Lower income neighbourhood patients with SCD VOC are associated with lower acuity triage scores and increased repeat ED visits. Neighbourhood income quintile subgroup analysis from the lowest-income (1) to the highest-income quintile (5) by A) ED visit rate; B) acuity triage CTAS score; C) repeat ED visits within 14- and 30-days; and F) any 1-year ED visit frequency between 0–3/yr, 4–9/yr or ≥10/yr; Error bars represent 95% CI.

### Higher acuity ED triage scores reflects improved downstream metrics in quality of care for adult patients with SCD VOC

Subgroup analyses within the adult SCD VOC cohort after adjusting for triage score showed that CTAS 2 scores (over CTAS 3 or 4) indeed captured patients with high ED visit frequencies (*Figure 5A*) and those with the longest admission length of stay (*Figure 5C*). While adult SCD VOC cases assigned CTAS 2 resulted in a mean −9.2 minutes (95% CI −13.9–-4.5) shorter wait time compared to general adult ED counterparts (*p* = 0.00014), CTAS 4 case comparisons demonstrated that adult SCD VOC cases waited significantly longer by 44.3 minutes (95% CI 18.4–70.2) instead (*p* < 0.0001) (CTAS 3 case comparisons were statistically equivocal) (*Figure 5B*). Only CTAS 2 (over CTAS 3 or 4) scores captured relatively equivocal repeat ED visit rates within 30-days for adult SCD VOC cases compared to general adult ED counterparts, whereas CTAS 3 or 4 scores reflected disproportionately higher repeat ED visit rates (*Figure 5E*). Higher acuity triage scores also correlated with a higher likelihood of hospital admission (*Figure 5D*) and fewer cases in the ≥10 visits/year bracket that otherwise barely registered in the reference cohorts (*Figure 5F*). This data altogether highlights the influence of SCD VOC-specific triaging parameters and demonstrates that higher acuity ED triage scores (i.e. CTAS 2 over CTAS 3 or 4) reflects improved downstream quality of care metrics for adult patients with SCD VOC.

**Figure 5:**
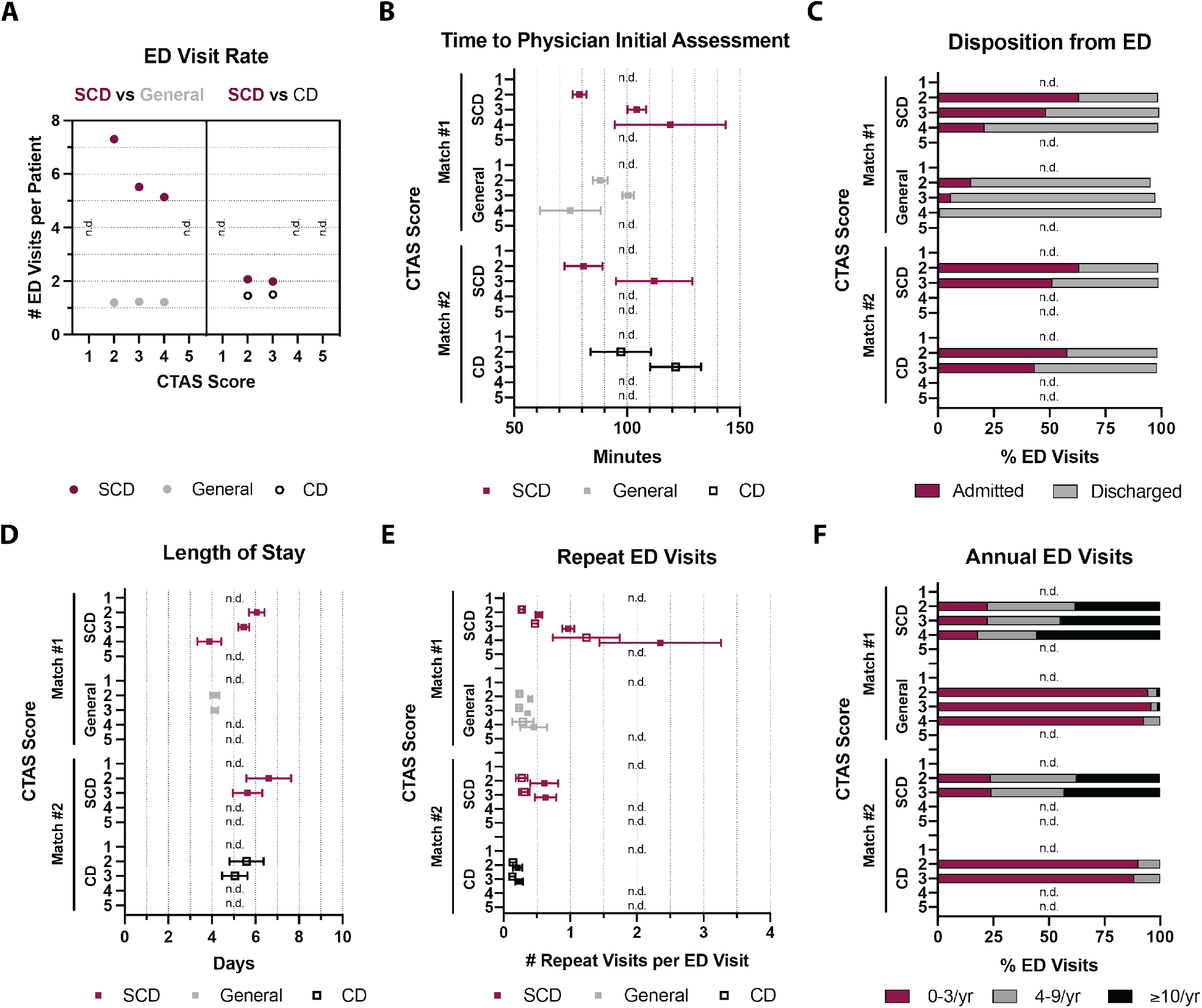
Higher acuity ED triage scores reflect improved downstream quality of care metrics for adults with SCD VOC. Correlated ED metrics and outcomes based on acuity triage CTAS score for A) ED visit rates; B) time to initial physician assessment; C) ED disposition; D) hospital admission length of stay; E) repeat ED visits within 14- or 30-days; and F) any 1-year ED visit frequency between 0–3/yr, 4–9/yr or ≥10/yr; Error bars represent 95% CI.

## Discussion

This population-based study of SCD VOC ED and inpatient quality of care for the last greater decade in Ontario, Canada has identified that patients with SCD VOC carry a significant burden of acute care needs, with a smaller patient subset whose high ED visit rates have distinctly increased over time. We found that adults, males and those from lower income quintile neighbourhoods with SCD VOC experienced lower quality of care metrics compared to their counterparts, with differences that were disproportionately larger than those found in the general ED population and among patients with Crohn’s disease. We also found that higher acuity ED triage scores – specifically CTAS 2 over CTAS 3 or 4 – reflected improved downstream quality of care metrics for adults with SCD VOC. Altogether, these findings raise concern that health inequities specific to patients with SCD VOC remain present within a universal and publicly funded healthcare system, and identifies opportunities for future work to close these gaps of care.

Our observations broadly highlight a need to identify and address determinants of health inequities in disease-specific settings. For example, why do adult SCD VOC ED wait times remain comparable to adults in the general ED population despite drastically different triage acuity scores? Why are there significantly different lag times for the same CTAS score between adult SCD VOC cases and those from the general ED population? And why does CTAS scoring differ in ways that correlate with neighbourhood income quintiles in SCD VOC but not in other patient populations? These striking differences in how patients with SCD VOC are triaged compared to two closely matched but diverse patient cohorts warrant further consideration of unaccounted factors that include but are not limited to patient and healthcare provider educational backgrounds, the role of structural racism, and/or other unaccounted biases within Canada’s healthcare system. CTAS criteria may also be better customized for use in disease-specific ways rather than a one-size-fits-all approach, adding to a growing recognition that future efforts for Canadian healthcare reform should focus on particular population health needs,^17^ especially that of marginalized and underserved patient communities that include SCD VOC.

Our study remains limited including our inability to further control for the aforementioned variable data within our health administrative databases, including the absence of clinical indicators of severity of presentation. Of note, use of the ICD-10-CA code D57.0 for sickle-cell anaemia with crisis captures other SCD overlapping presentations including acute chest syndrome and splenic sequestration, which may have contributed to some heterogeneity in our cohort establishment. Our identified care quality disparities represent static snapshots that cannot be strictly attributed to specific causes with full certainty. Finally, other important clinical metrics such as time to first dose analgesic and/or other medications would have further strengthened our analyses in the ED setting.

## Conclusions

Our work provides an evidence-based foundation to guide future quality improvement efforts for better SCD VOC care, shedding light on the particular needs of adults, males and those from lower income quintile neighbourhoods who experienced lower metrics in quality of care. We urge patient groups and multidisciplinary stakeholders to work collaboratively to improve decision-making in our health administrative systems (e.g. collecting educational and ethnic background data) and in how we can deliver better and more equitable care to patients (e.g. disease-specific adjustments to triage score criteria, quality improvement audits). Considering how long Canadians with SCD have reported on their varied healthcare experiences, we believe this work importantly addresses and delivers findings that are largely overdue.

## Data Availability

All data produced in the present study are available upon reasonable request to the authors.

## Acknowledgements

The authors thank Alexander Kopp for oversight of work completed by our data analysts, and Jessica Zung and Milena Hadsi-Tosev for statistical advice.

## Supplemental Content

### Cohort comparisons between patients with SCD VOC, the general ED population and patients with Crohn’s disease

Comparative analyses against matched reference cohorts from the general ED population and separately with patients diagnosed with Crohn’s disease showed that patients with SCD VOC were 5.5 to 13.0 times more likely to be triaged at a higher level of acuity (CTAS 1 or 2 over CTAS 3, 4 or 5) (*Figure 1D*). When further adjusted by including ED triage score into the matching criteria, the time to initial physician assessment for SCD VOC averaged 81.3 minutes (95% CI 79.2–83.3) to 93.9 minutes (95% CI 84.6–103.2), which were 9.1 (95% CI 6.3–11.8, *p* < 0.0001) to 14.9 minutes (95% CI 2.42–27.28, *p* = 0.019) faster than the reference cohorts (*Figure 1E*). Patients with SCD VOC stayed significantly longer in the ED from triage to disposition at means of 442.4 (95% CI 436.2–448.6) to 524.2 minutes (95% CI 499.8–548.6), which averaged 184.9 (95% CI 178.1–191.7, *p* < 0.0001) to 80.1 minutes (95% CI 48.5–111.7, *p* < 0.0001) longer than the reference cohorts. SCD VOC cases were 13.4 to 1.5 times more likely to be admitted to hospital than discharged from the ED compared to the reference cohorts (*Figure 1F*). Among those admitted, the length of hospital stay for SCD VOC averaged 5.2 (95% CI 4.98–5.32) to 6.1 days (95% CI 5.5–6.6), which was 1.2 days (95% CI 0.94 – 1.4, *p* < 0.0001) longer than the general ED population, and statistically equivocal compared to patients with Crohn’s disease (*p* = 0.16). Repeat ED visits after being discharged for SCD VOC within 30-days averaged 0.61 (95% CI 0.57–0.65) to 0.59 visits (95% CI 0.47–0.71), which was 0.28 (95% CI 0.24–0.32, *p* < 0.0001) to 0.37 visits (95% CI 0.25–0.49, *p* < 0.0001) more than the reference cohorts. Over any 1-year period, SCD VOC cases were 0.015 to 0.040, 21.5 to 4.7, and at least 55.3 times more likely to incur ED visits within a frequency of 1–3, 4–9, and ≥10 visits/year respectively compared to the reference cohorts (no patients with Crohn’s disease had ≥10 visits/year) (*Figure 1G*). Subgroup analyses between these various ED visit frequencies revealed no differences in triage scoring or ED disposition. However, higher ED visit frequencies in the SCD VOC cohort generally correlated with longer wait times, longer time between ED triage and disposition, and longer admission length of stay. These visits in the SCD VOC cohort were also more often distinct rather than repeats in nature compared to both reference cohorts (*Figure 1H*).

**eTable 1:** Demographic background of identified patients with SCD VOC as of December 31, 2018.

|  |  | **SCD VOC** |
| --- | --- | --- |
|  |  | *n* = 2,123 |
| **Age** | Mean ± SD | 24.78 ± 16.95 |
|  | Median (IQR) | 23 (11–35) |
|  | 0–4 (%) | 207 (9.8%) |
|  | 5–9 (%) | 269 (12.7%) |
|  | 10–18 (%) | 382 (18.0%) |
|  | 19–30 (%) | 568 (26.8%) |
|  | 31–40 (%) | 318 (15.0%) |
|  | 41–50 (%) | 197 (9.3%) |
|  | 51–60 (%) | 107 (5.0%) |
|  | 60+ (%) | 75 (3.5%) |
|  | <18 (%) | 821 (38.7%) |
|  | 18+ (%) | 1,302 (61.3%) |
| **Sex** | Male (%) | 1,005 (47.3%) |
|  | Female (%) | 1,118 (52.7%) |
| **Rurality** | Rural (%) | 12 (0.6%) |
|  | Urban (%) | 2,107 (99.2%) |
| **Neighbourhood Income** (Quintile) | 1 (Lowest) (%) | 797 (37.5%) |
|  | 2 (%) | 468 (22.0%) |
|  | 3 (%) | 455 (21.4%) |
|  | 4 (%) | 256 (12.1%) |
|  | 5 (Highest) (%) | 143 (6.7%) |
| **Residential Instability\*** (Quintile) | 1 (%) | 528 (24.9%) |
|  | 2 (%) | 280 (13.2%) |
|  | 3 (%) | 251 (11.8%) |
|  | 4 (%) | 391 (18.4%) |
|  | 5 (%) | 667 (31.4%) |
| **Material Deprivation\*** (Quintile) | 1 (%) | 219 (10.3%) |
|  | 2 (%) | 255 (12.0%) |
|  | 3 (%) | 334 (15.7%) |
|  | 4 (%) | 436 (20.5%) |
|  | 5 (%) | 873 (41.1%) |
| **Dependency\*** (Quintile) | 1 (%) | 1,009 (47.5%) |
|  | 2 (%) | 497 (23.4%) |
|  | 3 (%) | 267 (12.6%) |
|  | 4 (%) | 189 (8.9%) |
|  | 5 (%) | 155 (7.3%) |
| **Ethnic Concentration\*** (Quintile) | 1 (%) | 32 (1.5%) |
|  | 2 (%) | 76 (3.6%) |
|  | 3 (%) | 143 (6.7%) |
|  | 4 (%) | 412 (19.4%) |
|  | 5 (%) | 1,454 (68.5%) |

*\*Dimensions defined as per the 2016 Ontario Marginalization Index.*

**eTable 2:**
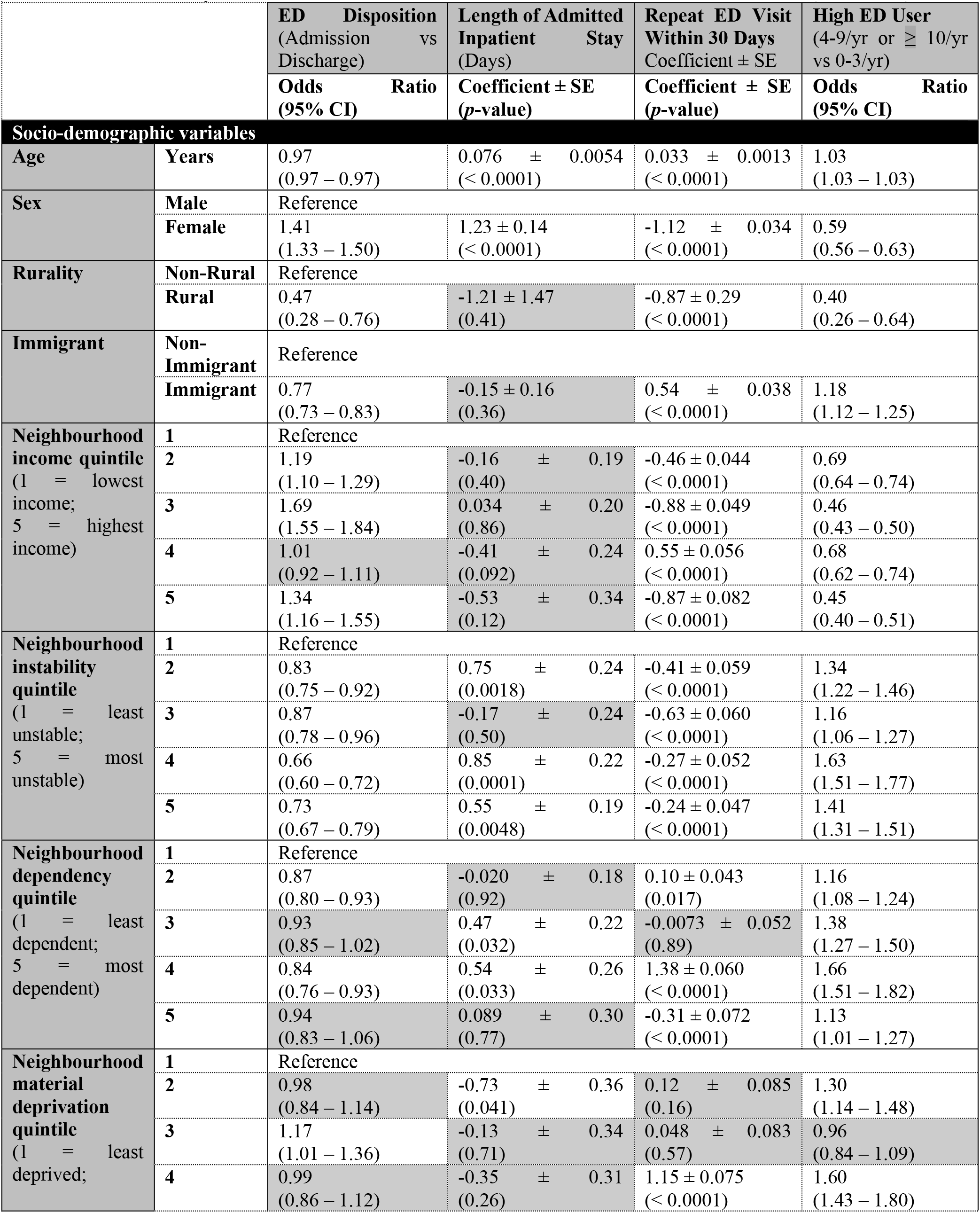
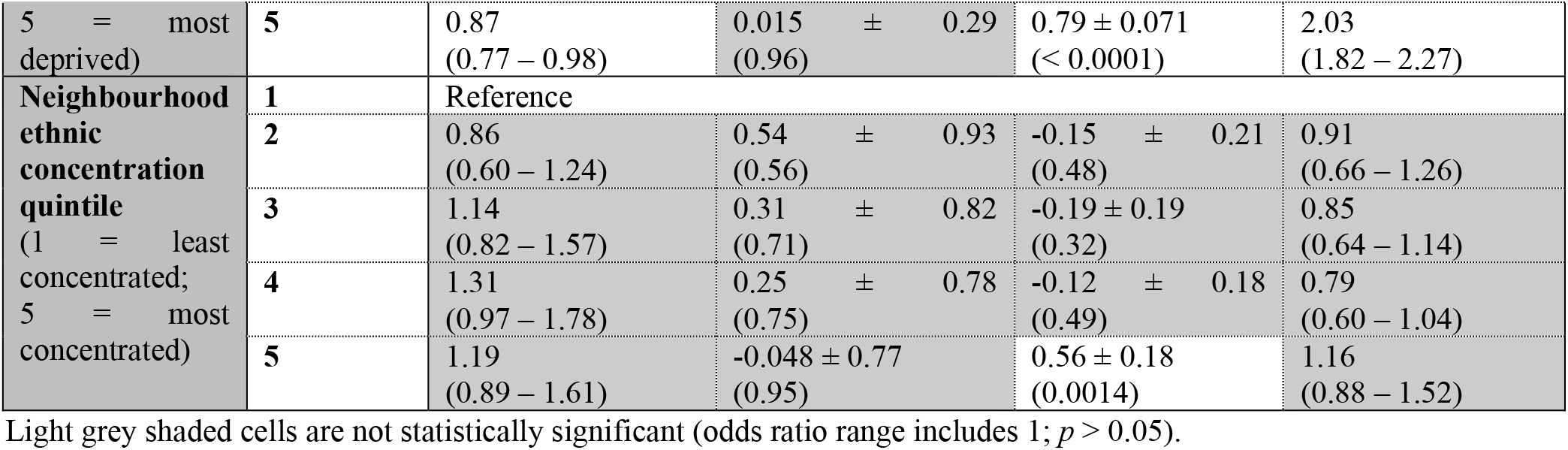
Bivariate analyses for ED-related clinical outcomes for SCD VOC in Ontario, Canada (2006–2018).

